# THE G-FORCE CONUNDRUM IN PRF GENERATION- MANAGEMENT OF A PROBLEM HIDDEN IN PLAIN SIGHT

**DOI:** 10.1101/2020.04.29.20084251

**Authors:** Kidambi Sneha, Jhansi Rani Ajmera, Rampalli Viswa Chandra

## Abstract

**Aim:** A force of 400g at 2700 RPM results in an optimum leucocyte and platelet-rich fibrin (L-PRF). Most of centrifuges with varying characteristics generate a g-force in excess of 700g at 2700 RPM. In this context, the study explores the effect of the original centrifugation protocol and a modified protocol tailor-made to lower the RPM to generate a g-force of ~400g on platelet concentration, clot size and growth factors release in L-PRF prepared in two different commercially available centrifuges.

**Materials and Methods:** 25 subjects each were assigned to the following groups; R_1_ and R_2_ where L-PRF was obtained from two laboratory swing-out centrifuges (Remi 8C® & Remi C854®, Mumbai, India) respectively. PRF was obtained from each subject within a group using two protocols; Original (O) protocol: conforming to the original centrifugation cycle (2700 RPM for 12 min) and Modified (M) protocol. Clot size, growth factor estimation and platelet counts were measured at 20, 40 and 60 mins from all the L-PRF clots.

**Results:** At the third time period (40–60min), there were no significant differences in clot sizes with the original protocol (*p=0.09*), but a highly significant difference was noticed with the modified protocol in both the centrifuges (*p=0.001*). Our results showed an increased concentration of VEGF and EGF with modified protocol than with original protocol with both the centrifuges (*p=0.001*). By the end of second and third time periods, more platelet concentration was observed with modified protocol than with the original protocol in both the centrifuges (*p=0.001*)

**Conclusion:** This study infers that the centrifuge type and RCF can affect the quality and quantity of cells and growth factors and an optimum relationship between g-force and RPM should be maintained in order to obtain L-PRF with adequate cell viability and optimum growth factor release.

## INTRODUCTION

PRF generation is a centrifugation-dependent process.^1^ Centrifuges work by putting supernatants in rotation around a fixed axis, thereby applying an accelerative force perpendicular to the axis. Relative centrifugal force (RCF; g-force) is the amount of accelerative force applied to a sample in a centrifuge, which is directly proportional to the revolutions per minute (RPM) a sample in a test-tube is subjected to.^2^ This resultant force causes the separation of various elements in the sample based on the individual weight of its elements and is the basis for blood separation techniques carried out by laboratory centrifuges.^1,2^

RCF (g) is measured in multiples of the standard acceleration due to gravity at the earth’s surface and is based on two specific variables which include the width/radius of the rotor and the speed of rotation (RPM).^2^ The radius of the centrifuge or rotor is as critical as the RPM in the process of producing a specific RCF.^2^ RPM and RCF are related by the formula RCF = 1.12*r* (RPM/1000) ^2^ where, r is the center of the centrifuge to tube end distance in millimeters.^1^ A force of 400g at 2700 RPM results in an optimum leucocyte- and platelet-rich fibrin (L-PRF).^3–7^ A myriad of centrifuges with different radii are designed and used in practice, which results in inappropriate architecture and cell content of L-PRF.^1,3–7^ RCF is an important parameter in the production of L-PRF and must be calculated for each centrifuge, especially if this parameter is not pre-set on the machine.^3^ Most often than not, running these centrifuges at 2700 RPM results in a g-force in an excess of 700g.^1–7^ At the same time, there is no provision for adjusting or changing the g-force through analog or digital means.^1,2^ If the center of the centrifuge to tube end distance in millimeters is known, by applying the abovementioned formula (*Figure 1*), the RPM can be altered to generate a force of 400g resulting in a L-PRF of better quality.^1–3^

**Figure 1:**
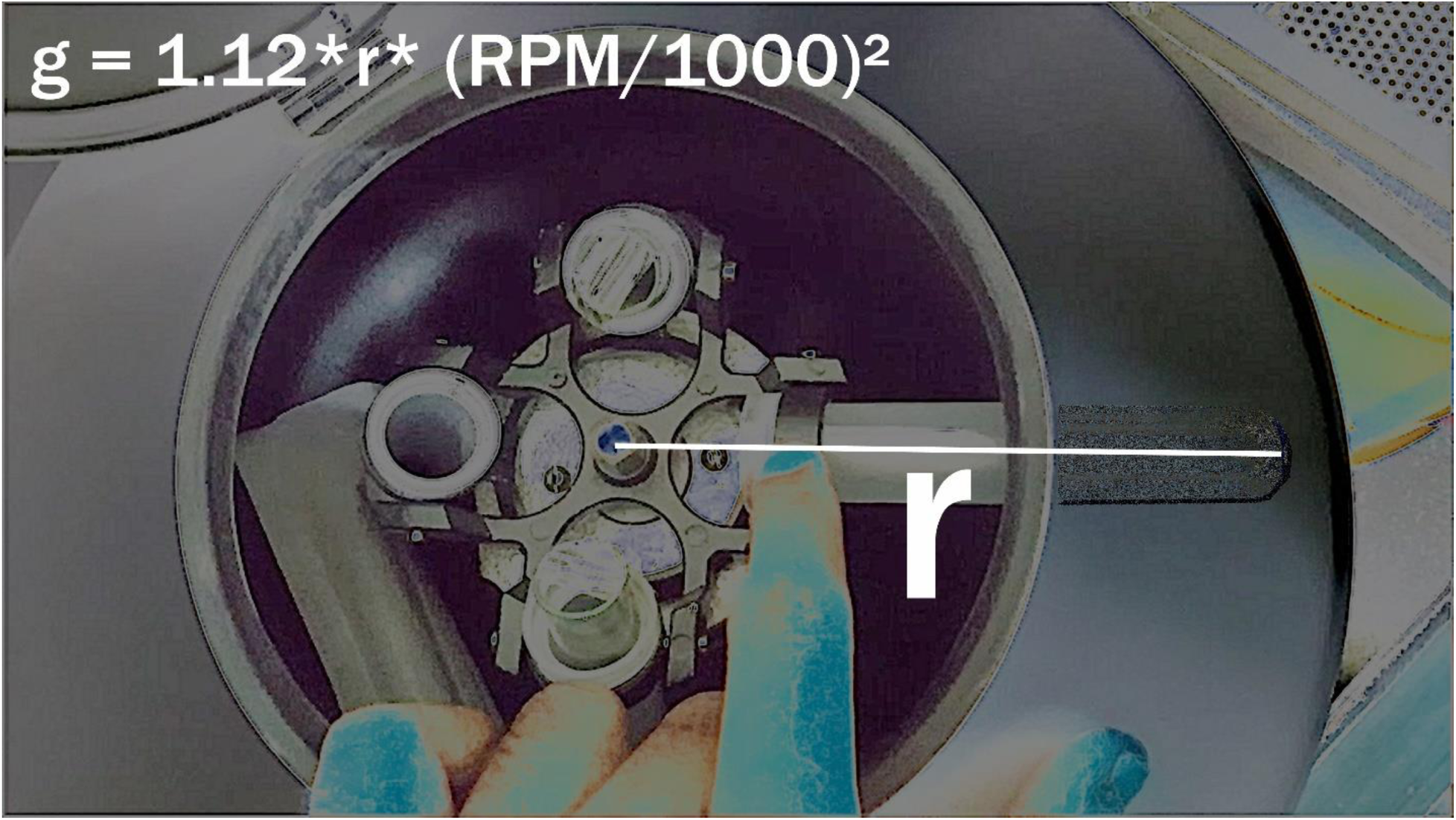
RPM and g-forces are related by the formula g=1.12*r* (RPM/1000) ^2^ where, r is the center of the centrifuge to tube end distance in millimeters.

This reduction in the g-force is extremely beneficial; Amable *et al*.,^4^ showed that changes in RCF significantly influence the platelet yield in platelet rich plasma when centrifuge time and temperature are kept constant.^4^ In a recent study,^5^ it was observed that the organization of the fibrin matrix and the release kinetics of growth factors are influenced by factors such as centrifugation time, g-force, type of rotor, model of the centrifuge as well as the type of tubes used for blood collection.^5^ When the g-force and RPM relationship is not appropriate, it results in the preparation of a clot of much smaller size, weaker biological significance and lower fibrin polymerization even when a stable centrifuge was used.^1–6^ This has a negative effect on the release of growth factors as well. Lowering the RPM controls and reduces the g-forces and results in an increase in cell number, platelets and growth factors such as VEGF and TGF.^7^ Therefore, this LSCC explains the importance of protecting the viability of cells and also the activation of various cells and growth factors.

In this context, the study explores the effect of the original centrifugation protocol and a modified protocol tailor-made to lower the RPM to generate a g-force of ~400g on platelet concentration, clot size and growth factors release in L-PRF prepared in two different commercially available centrifuges.

## MATERIALS AND METHODS

### Sample size and study population

To have an 80% chance (*β* error) of detecting a significant (two-sided 5% level) and a largest difference of 1mm in clot size between groups with a standard deviation of 1, 20 PRF clots per group were required. Accordingly, a total of 50 systemically healthy volunteers (mean age=26.87±8.76 years; 30 males), with no history of anti-coagulant intake were enrolled in the study.

#### Trial Design and Interventions

From all subjects, 50 ml of blood was drawn and was distributed into 8ml aliquots of six vacutainers (BD Vacutainer Plus Serum Tube®, Surgo, Totonto) after a clean venipuncture. 25 subjects each were assigned to the following groups; R_1_: participants from whom L-PRF was obtained from a laboratory swing-out centrifuge (Remi 8C®, Mumbai, India) and R_2_: where L-PRF was obtained from another laboratory swing-out Centrifuge with different characteristics (Remi C854®, Mumbai, India). PRF was obtained from a set of three aliquots from each subject within a group using two protocols; Original (O) protocol: conforming to the original centrifugation cycle (2700 RPM for 12 min) and Modified (M) protocol (*Figure 1*); The cycle was modified as follows. The G-force and RPM of a centrifuge are related by the formula RCF = 1.12*r* (RPM/1000) ^2^ where, r is the center of the centrifuge to tube end distance in millimeters.^1^ The “r” values of both the centrifuges were measured. The RPM required to generate 400 g of RCF in a 125-mm tube of R_1_ was calculated (RPM = 1690) and in a tube of 130 mm (R_2_; 1650 RPM) were calculated. The RPMs were rounded to PRFs were obtained at 1700 RPM in both the centrifuges.

### Outcomes

#### Clot size

After centrifugation, the L-PRF clot was removed from the test tube and a smooth spatula was used to gently release the red clot from the buffy coat. The clots were measured in length at breadth using a Vernier calipers at 20, 40 and 60 mins (Times A, B & C) respectively. The average of the lengths (V) and the breadths (B) was considered to be the clot size (*Figure 2*).

**Figure 2:**
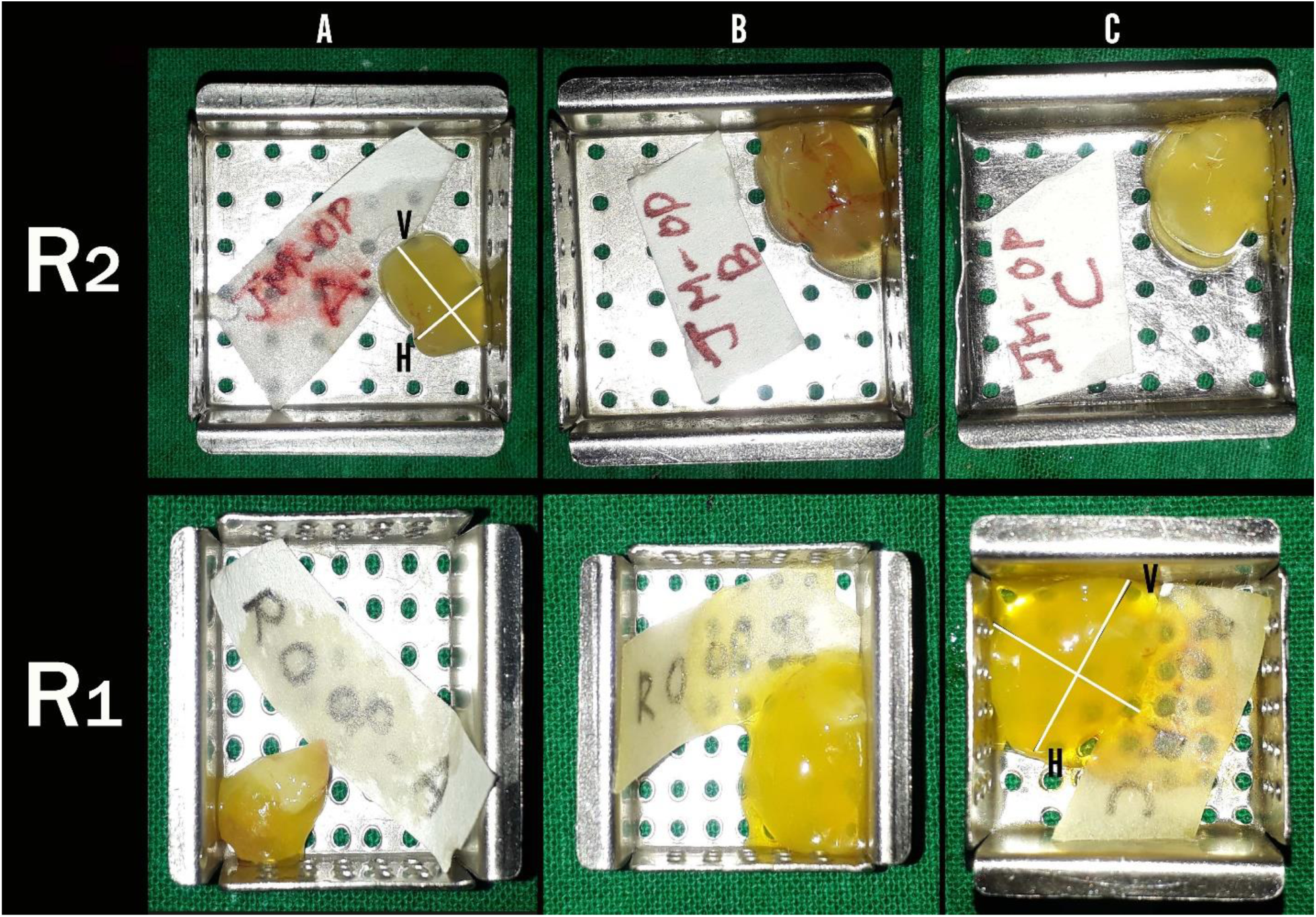
This figure depicts the general trend in clot sizes generated by the two centrifuges R_1_ and R_2_ at different time-frames. It can be observed that the clots formed with R_1_ were larger in size than that of R_2_. ‘V’ and ‘H’ are the vertical and horizontal dimensions of the clot respectively.

#### Growth Factor Estimation

After 20 mins, PRF clots were retrieved from the vacutainers and RBC layer was detached and discarded. Four PRF clots were transferred into sterile tubes and were agitated gently for 5 minutes (Agitaser®, Barcelona, Spain). The clot was then minced in a 7 mL tissue grinder (Tenbroeck®, Bangalore, India) to obtain a releasate which was measured and the releasate returned into the tube. The releasates were immediately centrifuged at 10,000g for 15 minutes (Microfuge22R®, Beckman Coulter, Fullerton, CA) to pellet out any residual blood cells, and supernatants were frozen at −80°C till determination of the growth factors (vascular endothelial growth factor (VEGF) and Epidermal growth factor (EGF)). Two commercially available ELISA kits were used to measure VEGF^8^ (PicoKine™, Bosterbio, Pleasanton, USA) and EGF^9^ (Human EGF ELISA Kit®, Origene, Rockville, USA) levels respectively as per the instructions of the manufacturers.^8,9^

#### Platelet count

After at 20, 40 and 60 mins (Times A, B & C), RBC layer was removed and the clots were compressed gently to remove excess fluid; the remaining white PRF matrix was fixed in 10% formalin for 24 h and dehydrated in a series of ethanol solutions (starting at 70% and reaching 100%) for use as histological specimens. H and E stained sections were obtained and each slide, ten regions of interest (ROIs; Figure 3) per slide were imaged (Olympus BX53® microscope, DSS Group, New Delhi, India) at 40X magnification. Platelets were counted as per the technique of Li.^10^

**Figure 3:**
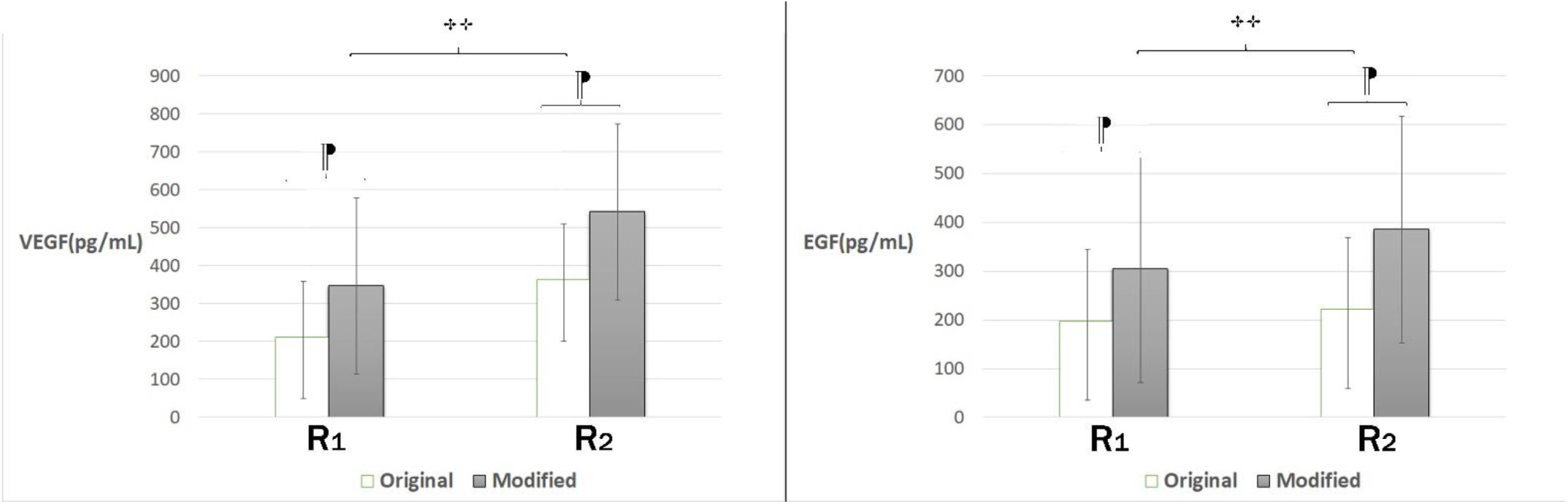
R_2_ showed a highly significant release of VEGF and EGF (pg/mL) over R_1_ regardless of the protocol (*p=0.001*\*\*). Our results showed an increased concentration of VEGF and EGF with modified protocol than with original protocol in both the centrifuges (*p=0.001*¶).

**Figure 4:**
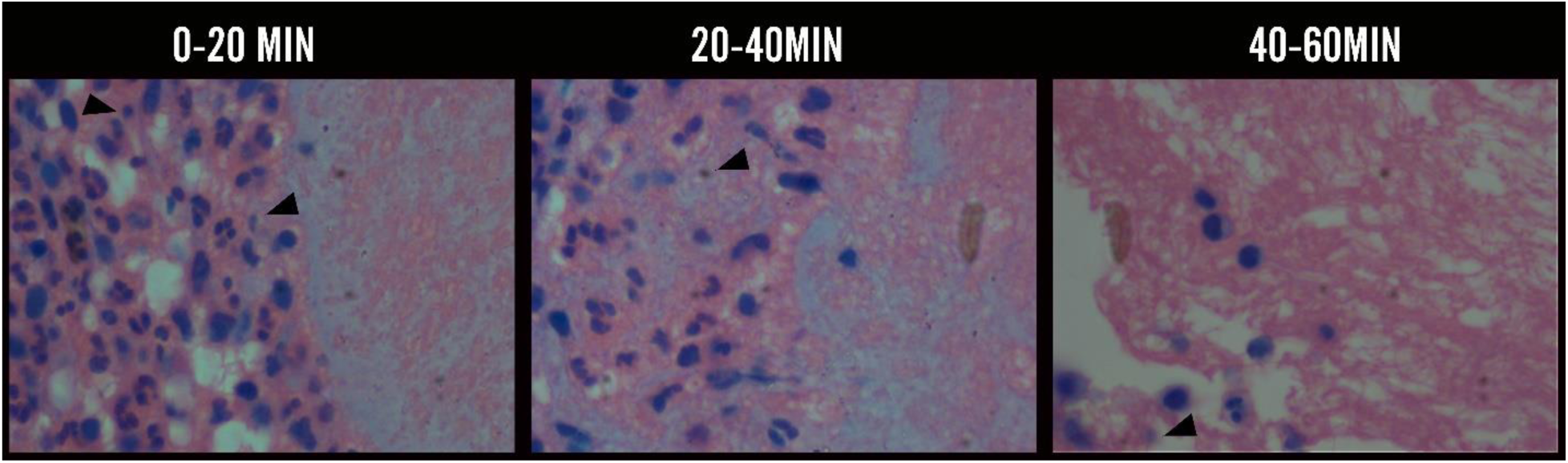
This picture depicts the general trend in platelet concentration in three PRF specimens taken at three different time periods in both the centrifuges run with two protocols. Given picture shows the presence of numerous white blood cells, lumen and platelets (small sized cells present adjacent to the WBC, *arrow*). There was a gradual decrease in the platelet concentration from baseline to 60 min with the original protocol in both centrifuges (*p=0.001*).

#### Statistical Analysis

Data was analyzed by using Prism8^®^ (GraphPad Software, La Jolla, USA). Intragroup comparison was performed by using ANOVA followed by multiple comparisons using Bonferroni correction. One-way ANOVA followed by the post hoc test was used for intragroup and intergroup comparisons. A p≤0.001 was considered as highly significant, p≤0.05 as significant and p>0.05 as non-significant.

## RESULTS

### Morphological analysis

*Figure 2* shows the difference in clot sizes produced by two different centrifuges with two protocols (original(O) and modified (M)). At the first time period (0–20min), there were no significant differences in clot sizes with both protocols in the two centrifuges (O: *p=0.08;* M: *p=0.3*). Whereas, at the second time period (20–40min), the original protocol showed significant (*p=0.03*) to highly significant differences (*p=0.001*) in clot size (R_1_-O: 3.43±1.21; R_2_-O: 3.78±1.69; R_1_-M: 3.18±1.92; R_2_-M: 3.62±2.01) over the modified protocol in both the centrifuges. At the third time period (40–60min), there were no significant differences in clot sizes with the original protocol (R_1_-O: 4.12±2.01; R_2_-O: 3.99±1.90) (*p=0.09*), but a highly significant difference was noticed with the modified protocol in both the centrifuges (R_1_-M: 4.89±1.79; R_2_-M: 3.79±1.22 (*p=0.001*) (*Table 1*).

**Table 1:**
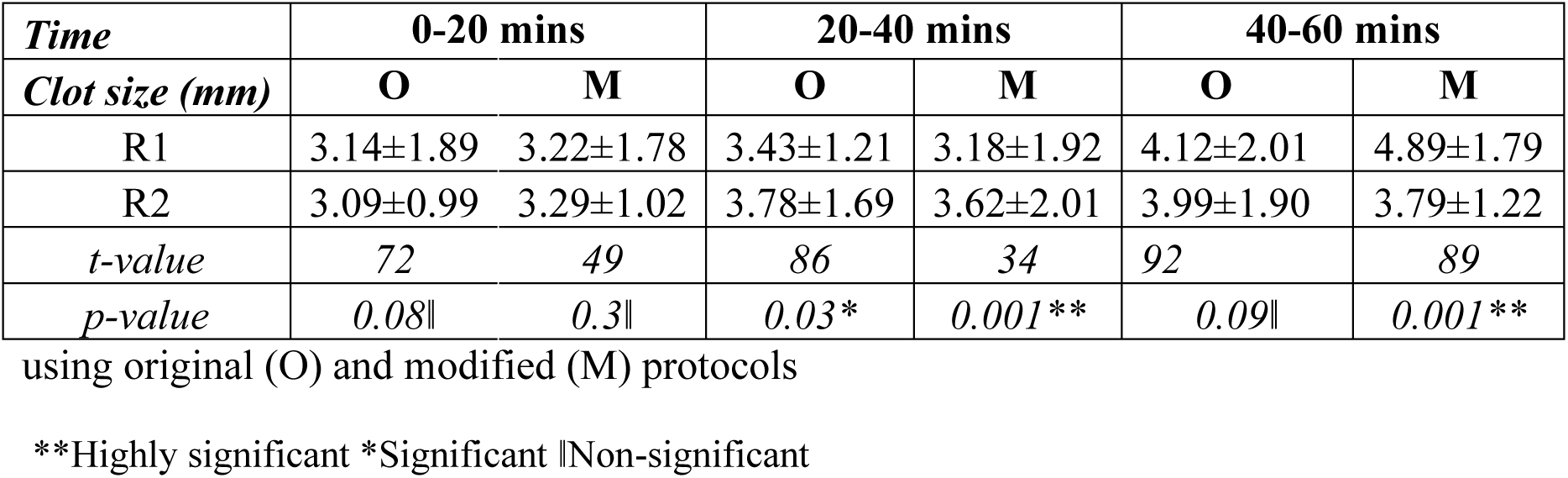
Comparison of clot sizes at three different time periods with two different centrifuges

### Growth factor release and Platelet counts

The release of VEGF (pg/mL) in R_1_ and R_2_ centrifuges showed a highly significant difference with the two protocols (R_1_-O: 212±146, R_1_-M: 347±163; R_2_-O: 363±232; R_2_-M: 542±303) (*p=0.001*). The release of EGF (pg/mL) also showed a highly significant difference with original (R_1_-O: 198±96; R_2_-O: 222±142) and modified protocols (R_1_-M 304±122; R_2_-M 385±212) in both the centrifuges (*p=0.001*). Our results showed an increased concentration of VEGF with modified protocol than with original protocol with both the centrifuges (*p=0.001*). An increased concentration of EGF was observed with modified protocol when compared to original protocol with both the centrifuges (*p=0.001*) (*Table 2*).

**Table 2:** Group and sub group analysis of growth factors’ and platelet concentration at different time periods with two centrifuges using original (O) and modified (M) protocols.

| Group and Sub-group analysis |  | Sub-group analysis |  |  | *t value* | *p value* |
| --- | --- | --- | --- | --- | --- | --- |
|  |  | Protocol | R1 | R2 |  |  |
| Group Analysis | VEGF (pg/mL) | O | 212±146 | 363±232 | 232 | 0.001** |
|  |  | M | 347±163 | 542±303 | 302 | 0.001** |
|  |  | *t value* | 132 | 262 |  |  |
|  |  | *p value* | 0.001** | 0.001** |  |  |
|  | EGF (pg/mL) | O | 198±96 | 222±142 | 64 | 0.001** |
|  |  | M | 304±122 | 385±212 | 162 | 0.001** |
|  |  | *t value* | 176 | 198 |  |  |
|  |  | *p value* | 0.001** | 0.001** |  |  |
|  | Platelets (*10<sup>8</sup>) at 0-20 mins | O | 4.5±0.82 | 3.4±0.85 | 92 | 0.001** |
|  |  | M | 3.5±0.59 | 1.8±0.88 | 12 | 0.001** |
|  |  | *t value* | 64 | 82 |  |  |
|  |  | *p value* | 0.001** | 0.001** |  |  |
|  | Platelets (*10<sup>8</sup>) at 20-40 mins | O | 1.15±0.92 | 2.15±0.93 | 24 | 0.001** |
|  |  | M | 2.7±0.23 | 2.85±9.22 | 43 | 0.036* |
|  |  | *t value* | 98 | 67 |  |  |
|  |  | *p value* | 0.001** | 0.04* |  |  |
|  | Platelets (*10<sup>8</sup>) at 40-60 mins | O | 1.1±0.65 | 0.6±0.22 | 67 | 0.001** |
|  |  | M | 2.5±1.1 | 3.15±2.01 | 34 | 0.001** |
|  |  | *t value* | 21 | 109 |  |  |
|  |  | *p value* | 0.001** | 0.001** |  |  |

\*\*Highly significant \*Significant

For platelet counts (*10^8^), in the first time period (0–20min), both the centrifuges showed a highly significant difference between the two protocols, with more concentration of platelets observed in original protocol (R_1_-O: 4.5±0.82; R_2_-O: 4.5±0.82) than with modified protocol (R_1_-M: 3.5±0.59; R_2_-M: 1.8±0.88) (*p=0.001*). But in the second time period (20–40min), R_1_ centrifuge showed highly significant difference between original and modified protocols where, more platelet concentrations were observed with modified protocol (R_1_-M: 2.7±0.23) than with the original protocol (R_1_-O: 1.15±0.92) (*p=0.001*); also, a significant difference was noticed with R_2_ centrifuge between the two protocols (R_2_-O: 2.15±0.93; R_2_-M: 2.85±9.22) (*p=0.04*). Also, in the third time period, both the centrifuges showed a highly significant difference between both the protocols (*p=0.001*). Here, an increase in platelet count was observed in R_2_ centrifuge with the modified protocol (R_2_-M: 2.85±9.22). In the first time period, there was an increased platelet concentration observed with original protocol than with modified protocol. But in the second and third time periods, more platelet concentration was observed with modified protocol than with the original protocol with both the centrifuges (*p=0.001*) (*Table 2*).

## DISCUSSION

The present study aimed to compare the biological integrity of L-PRF prepared by two centrifuges using two types of protocols and determined its influence on clot size, growth factor concentration (VEGF and EGF) and on platelet concentration. This study highlighted the importance of RPM-G force relationship in obtaining accurate PRF clots as it affects the cell viability and activation of the cell contents.

When g-forces were lowered for the preparation of L-PRF in both the centrifuges, R_1_ (4.89±1.79 mm), a much powerful centrifuge than R_2_ (3.79±1.22 mm), resulted in larger clot comparatively, as an appropriate RPM-g force relationship was maintained. The smaller clot size formed from R_2_ centrifuge might be because of a g-force insufficient for the proper and complete separation of blood constituents. A recent study^6^ has observed that, when an inappropriate g-force was used with a stable centrifuge, it resulted in a clot of much smaller size, weaker biological signature and lower fibrin polymerization. As centrifugation speed is decreased, the relative separation in layers of PRF is minimized and PRF clots formed are also smaller in size.^7^

The continuous release of growth factors is one of the main objectives justifying the use of platelet concentrates in regenerative medicine.^10–13^ When g-forces were lowered, it was observed that the concentration of VEGF (347±163 pg/mL) and EGF (304±122 pg/mL) increased when compared to the original protocol (VEGF:212±146 pg/mL) (EGF: 198±96 pg/mL) (*p=0.001*). El Bagdadi et al.,^11^ studied the platelet distribution pattern and growth factor release by preparing PRF at different relative centrifugation forces (RCF) and centrifugation times and observed that the reduction of RCF, lead to increased growth factor release in leukocytes and platelets within the solid PRF matrices.^11,12^ A study conducted by de Oliveira et al.,^5^ concluded that the smallest g-forces were more promising with the shape of the fibrin network in the PRF and also favored the release of VEGF from platelet granule store, culminating to the highest concentration of the growth factor which was observed up to 7 days.^5^

Kobayashi^13^ indicated that low-speed centrifugation concept favored an increase in growth factor release from PRF clots which in turn may directly influence the tissue regeneration by increasing fibroblast migration, proliferation and collagen mRNA levels. Since high centrifugation forces are known to shift the cell population to the bottom of collection tubes, it was hypothesized that by reducing centrifugation g-force, an increase in leukocyte numbers may be achieved within the PRF matrix.^13,14^ The g-force tends to change based on the location at which it is calculated along the test-tube, but it has been proved that the g-force calculated at the end of the centrifugation tube does not subject to change owing to the centrifugation time, even when centrifuged at the exact same speed.^15^

The histological report of the present study showed a gradual decrease in the platelet concentration from baseline to 60 min with the original protocol in both centrifuges. A decrease in the platelet concentration was also observed with modified protocol in centrifuge R_1_. But, interestingly, there was an increase in the platelet concentration with modified protocol in R_2_ (from 1.8±0.88 to 3.15±2.01*10^8^). It was also observed that the clots prepared with R_1_ centrifuge displayed cells with stable shape and size compared to that of R_2_. The main difference in platelet distribution might have occurred due to the difference in centrifugation speed.

This decrease in the platelet concentration can be explained by an *in vitro* clot examination study at different time intervals. The study showed that the platelet membrane disintegration occurred as the clot formation progressed.^16^ Initially, platelets in plasma were rounded with continuous limiting membrane, but gradually there was disruption of the limiting membrane and change in the shape of platelets followed by small platelet aggregate formation.

No individual intact platelets or any complex aggregates were observed with increase in time. Gradually, the clot became denser, the mass consisted only of fibrin and a few poorly defined membrane remnants.^16^ However it is difficult to assess the number of platelets that were totally disrupted during the PRF preparation.^17^ Contact with foreign surfaces, irrespective of their nature causes quick agglutination and lysis of thrombocytes, which might have also resulted in the reduction of the platelet count in the PRF.^16^

The quality of the clot started to deteriorate by the end of the third time period (40-60mins). Dohan Ehrenfest et al.,^18^ stated that the clot slowly starts to sink into the tube after centrifugation and merges with the red blood cell base, leading to an unusable material loaded with red blood cells with weak mechanical properties. Su et al.,^19^ proposed PRF membrane to be used immediately after formation and the use of a nonabsorbable impermeable sterile material and a sterile curvette to squeeze the PRF clot to maximize release of growth factors to the surgical site.^19^

This study infers that the centrifuge type and RCF can affect the quality and quantity of cells and growth factors. Although, this study did not evaluate L-PRF in all the designs of centrifuges, the inferences of this study can be applied to the other designs and can be standardized accordingly. Apart from speed, the types of tubes may also influence the platelet distribution,^20^ but however, the tube type has not been included as a parameter in the present study. Even if this difference does not influence the initial growth factor content, it may influence the nature of growth factor retention and release.^20^

To conclude, this study establishes the principle that when different designs of centrifuges are used, an optimum relationship between g-force and RPM should be maintained in order to obtain L-PRF with adequate cell viability and optimum growth factor release. This study also offers an opinion that the prepared PRF should be used in surgical defects immediately after its preparation as delaying would result in disintegration of platelets which may in turn affect the growth factor release.

## Data Availability

All data is available on request and enquiries can be sent to

